# Deep learning-based polygenic scores enhance generalizability of psychiatric disorders prediction

**DOI:** 10.1101/2025.05.05.25326794

**Authors:** Leonardo Cobuccio, Arnor I. Sigurdsson, Kajsa-Lotta Georgii Hellberg, Morten Dybdahl Krebs, Jonas Meisner, iPSYCH Study Consortium, Thomas Werge, Michael E. Benros, Andrew J. Schork, Simon Rasmussen

## Abstract

Polygenic scores (PGSs) have emerged as promising tools for predicting complex traits from genetic data, however, their predictive performance for psychiatric disorders remains limited and the added value of deep learning (DL) over linear models is underexplored. In this study, we compared our DL model, Genome-Local-Net (GLN), with the linear model bigstatsr in predicting five psychiatric disorders—ADHD, ASD, BIP, MDD, and SCZ—using individual-level genotype data. We further assessed whether combining these *internal* (individual-based) PGSs with *external* (GWAS-derived) PGSs and family genetic risk scores (FGRSs) could improve prediction additively or synergistically. While GLN and bigstatsr performed similarly in-sample, GLN showed better generalization on an out-of-sample replication set for ADHD, ASD, and MDD, with an average AUROC gain of 0.026. Integrating *internal*, *external*, and family-based scores significantly improved ADHD prediction, though DL-based integration provided no consistent advantage over logistic models. These findings suggest that while DL may enhance generalizability for specific psychiatric traits, linear models remain competitive and effective for genetic risk prediction.

## INTRODUCTION

The ability to predict disorders from genetic data is a focal point of genomic research, as it has the potential to advance early diagnosis and personalized medicine, while providing deeper insights into the genetic basis of medical conditions (Kullo et al., 2022; Torkamani et al., 2018). Research into genetic-based prediction has primarily focused on polygenic scores (PGSs), which can be divided into two types based on how SNP effect sizes are derived: *external* PGSs use effect sizes of SNPs estimated from external GWAS summary statistics, while *internal* PGSs estimate the SNP effects directly from individual-level data. PGSs have been found to have significant predictive accuracy for various medical conditions, including diabetes (Luckett et al., 2023), cardiovascular diseases (Mars et al., 2020), inflammatory bowel disease and breast cancer (Khera et al., 2018). However, even though psychiatry pioneered the development of PGSs (Purcell et al., 2009), the predictive performance of PGSs for mental disorders falls short of achieving similar levels of accuracy compared to some of the non-psychiatric conditions (Andreassen et al., 2023). For instance, current PGSs explain ∼11% of liability for schizophrenia and ∼4% for major depression, corresponding to AUCs of 0.74 and 0.60, respectively, values below thresholds typically considered clinically actionable (Murray et al., 2021). This likely reflects the complex and heterogeneous nature of psychiatric disorders, along with sample sizes that remain underpowered, highlighting the need for methods better suited to these challenges.

Many software packages have been developed to compute both *external* PGSs, like PRSice (Choi & O’Reilly, 2019), PRS-CS (Ge et al., 2019), SBayesR (Lloyd-Jones et al., 2019), LDpred2 (Privé et al., 2021), SDPR (Zhou & Zhao, 2021), and *internal* PGSs, like BOLT-LMM (Loh et al., 2015), bigstatsr (Privé et al., 2018, 2020), snpnet (Li et al., 2021; Qian et al., 2020), but they generally assume additive contributions from independent variants (Schwarzerova et al., 2024). The hypothesis of whether there is substantial contribution of non-linear and interactions effects between genetic variants for complex traits remains an intensely debated topic (Hemani et al., 2021; Hill et al., 2005; Sheppard et al., 2021; Tang et al., 2022). Nonetheless, exploring these effects remains a crucial area of investigation in understanding the genetic architecture of complex traits and could potentially lead to models with higher predictive performance. Recently developed non-linear Machine Learning (ML) and Deep Learning (DL) based PGSs were found to achieve at least similar performance to linear models, and outperforming them in a few specific cases such as autoimmune diseases (Ohta et al., 2024; Sigurdsson et al., 2023), Alzheimer’s (Hermes et al., 2023), inflammatory bowel disease (Klau et al., 2023; Peng et al., 2024), and breast cancer (Badré et al., 2021). Different studies have also developed DL-based predictors of psychiatric disorders from individual data, but they generally have low sample sizes and focused on maximum few thousands of SNPs as predictors (Bracher-Smith et al., 2021). Moreover, it is unclear how DL-based PGSs perform for psychiatric disorders compared to linear models, as many published studies were found to suffer from methodological issues such as overfitting, unfair comparisons, and data leakage, which undermine their validity and reliability (Bracher-Smith et al., 2021).

For complex disorders, a single *internal* or *external* PGS will never provide a complete picture of risk. Biomarkers, health traits, health states, constitutional factors, and family history, among other factors, will contribute and should be considered in a combined and comprehensive way (Schork et al., 2018). Among these, family history of disease is a common predictor that was shown, for many disorders, to be complementary to PGSs (Dybdahl Krebs et al., 2023; Hujoel et al., 2022; Mars et al., 2022). Recent work has emphasized how Family Genetic Risk Scores (FGRSs), which combine information from multiple relatives into a single score (Kendler et al., 2021), can significantly enhance prediction beyond PGSs alone. In major depressive disorder (MDD), for instance, both PGS and FGRS individually achieved modest classification performance (AUC ≈ 0.57–0.60), but their combination yielded a notable improvement (AUROC = 0.630 in iPSYCH1 and 0.608 in iPSYCH2), highlighting their complementary value for risk stratification (Dybdahl Krebs et al., 2024). Other studies have explored predicting psychiatric disorders by combining multiple external PGSs (Albiñana et al., 2023; Krapohl et al., 2018), *internal* and *external* PGSs (Albiñana et al., 2021), and they all have shown improved predictive performance compared to single feature models. However, the integration of *internal* and *external* PGSs with FGRSs, and further exploration of potential interactions among scores is currently understudied and could potentially provide a more powerful and comprehensive approach to genetic risk prediction in psychiatric disorders. In particular, DL approaches may offer an effective strategy for achieving this goal. In this scenario, we recently developed EIR, a DL framework designed to enhance individual-level PGS prediction by introducing a novel neural network architecture called Genome-Local-Net (GLN) (Sigurdsson et al., 2023). When we applied the GLN model from EIR to the UKBB dataset for predicting diagnoses and biomarker levels, we found it to outperform linear models in some cases (Sigurdsson et al., 2022, 2023, 2024). However, this model has never been applied on large scale psychiatric disorders cohorts.

Here we explored the use of the DL architecture GLN from the EIR framework (Sigurdsson et al., 2023) for predicting 5 psychiatric disorders: Attention Deficit Hyperactivity Disorder (ADHD), Autism Spectrum Disorder (ASD), Bipolar Disorder (BIP), Major Depressive Disorder (MDD), and Schizophrenia (SCZ), using large-scale genotype data from the Integrative Psychiatric Research Consortium (iPSYCH) (Pedersen et al., 2018; Bybjerg-Grauholm et al., 2020), and compared it to a state-of-the-field linear model, bigstatsr (Privé et al., 2018). We also explored the potential of combining GLN with scores from bigstatsr, *external* PGSs and FGRSs in both logistic regression- and DL-based integration approaches. We found that GLN and bigstatsr had similar performance in predicting the 5 psychiatric diagnoses on the test set, but GLN significantly outperformed and had better generalization performance than bigstatsr on the out-of-sample replication set for ADHD, ASD, and MDD. The integration of GLN and bigstatsr did not provide additional predictive performance, but combining GLN or bigstatsr with *external* PGS and FGRS did significantly improved prediction performance in terms of Area Under the Receiver Operating Curve (AUROC) for ADHD, suggesting that combining various types of genetic risk scores – both *internal* and *external* - could enhance the ability to predict some complex psychiatric phenotypes.

## RESULTS

### Study overview

In this study we leveraged the Danish population-based Integrative Psychiatric Research Consortium (iPSYCH) case-cohort dataset, which contains genotype data from individuals born in Denmark between 1981 and 2008 and followed up until 2016. The iPSYCH cohort consists of two waves of data: iPSYCH1 and iPSYCH2, which were genotyped on two different SNP arrays (see Methods). Taken together, the two waves contain data on 137,251 individuals, of which 88,831 were diagnosed with at least one of 5 psychiatric disorders: Attention Deficit Hyperactivity Disorder (ADHD), Autism Spectrum Disorder (ASD), Bipolar Disorder (BIP), Major Depressive Disorder (MDD), and Schizophrenia (SCZ). For controls, 48,420 individuals were randomly drawn from the entire population. The specific numbers of cases and controls in the two waves that we analyzed in this study are presented in **Figure 1C**.

**Figure 1.**
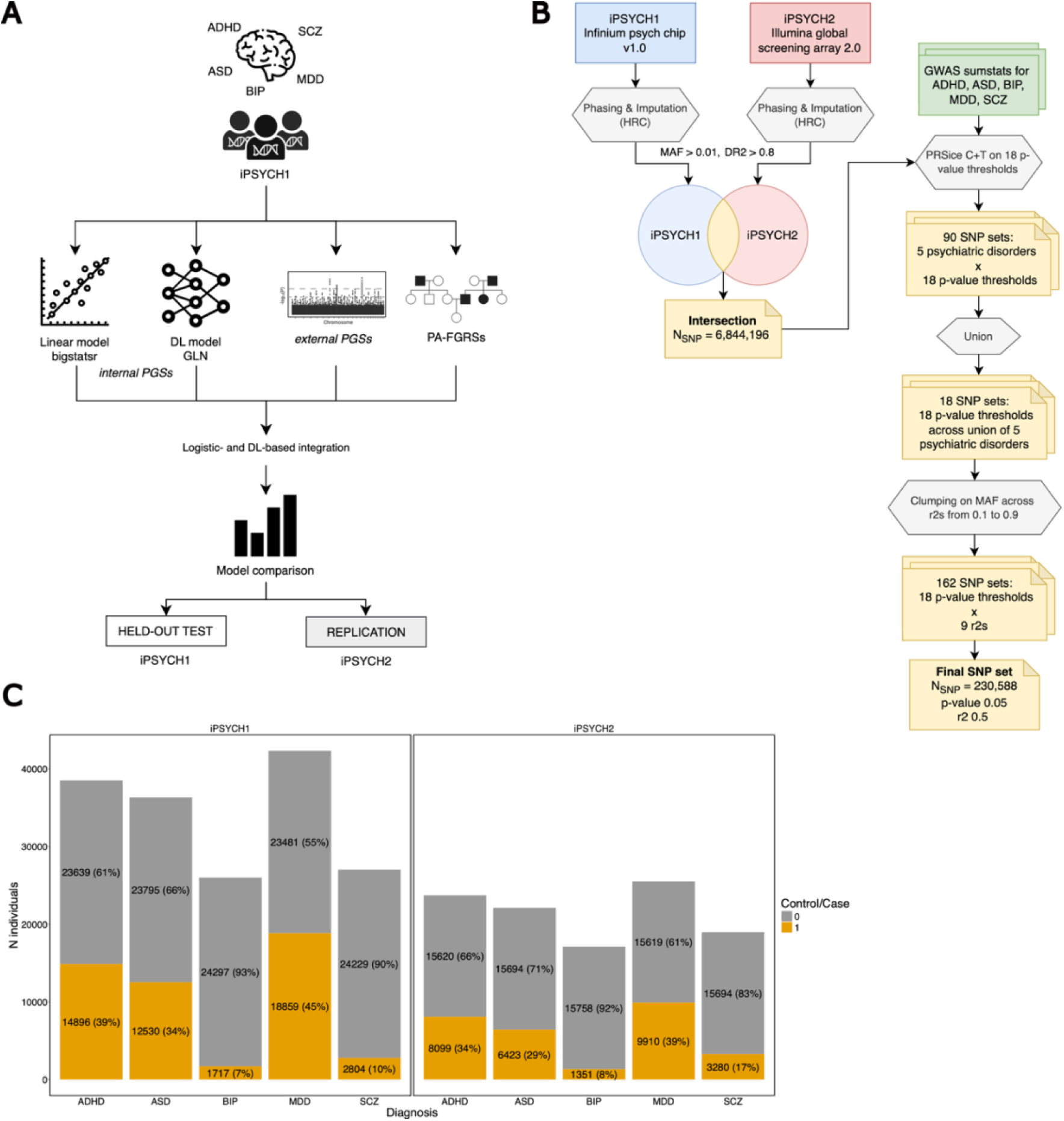
Overview of the study. **A)** Overview of the workflow followed in this study. After splitting the data into training and test, we used the data of the individuals from the iPSYCH1 cohort to train bigstatsr (a linear model) and GLN (a DL model) to compute *internal* PGSs for 5 different psychiatric disorders – ADHD, ASD, BIP, MDD, SCZ. On these individuals we also computed *external* PGSs and PA-FGRSs. We compared the performance of the single scores and explored their integration with a regression- and a DL-based approach, both on the held-out test of iPSYCH1 and on the entirety of the iPSYCH2 as out-of-sample replication set. **B)** Workflow of the strategy we used to select the final set of SNPs on which we computed the PGSs. After filtering for MAF > 0.01 and imputation score > 0.8, we extracted the intersection of the SNPs across the two cohorts iPSYCH1 and iPSYCH2. Using external GWAS summary statistics, we ran Clumping and Thresholding (C+T) with PRSice at different p-value thresholds, restricting the analysis on the intersected SNP set. This resulted in the creation of 90 SNP sets. After taking the union of these sets across each p-value threshold, we then performed clumping on MAF across a range of r2 values. Finally, we arbitrarily chose the final set of SNPs as the one created with parameters p-value 0.05 and r2 0.5, which consisted of 230,588 external GWAS-informed SNPs. See methods for more details. **C)** Distribution of the cases and controls across the 5 psychiatric disorders in the two iPSYCH cohorts 1 and 2.

A schematic overview of the study is presented in **Figure 1A**. For each of the 5 psychiatric disorders ADHD, ASD, BIP, MDD, SCZ, we used the genotype data of the individuals from the training sets of the iPSYCH1 cohort to compute two *internal* Polygenic Scores (PGSs): a linear one using bigstatsr, a state-of-the-art linear model (Privé et al., 2018, 2020), and our DL-based GLN model from the EIR framework (Sigurdsson et al., 2023). We further computed *external* PGSs with SBayesR and Pearson-Aitken Family Genetic Risk Scores (Dybdahl Krebs et al., 2024) (PA-FGRSs, hereinafter referred to as FGRSs for simplicity). We first compared the performances of the two *internal* PGSs, separately, and then assessed their contribution within integrated models that combined *external* PGSs and FGRSs, using both a logistic model and a DL-based approach. We evaluated the performances on a held-out test set from the iPSYCH1 cohort, and used the entirety of the iPSYCH2 cohort as an out-of-sample replication set. All PGS were computed using the same genotype data, a set of ∼230k GWAS-informed SNPs that were in common between the two iPSYCH (see Methods for more details and **Figure 1B** for a schematic overview).

### Hyperparameter tuning improves DL model performance for predicting psychiatric disorders

To optimize the DL model GLN from the EIR framework on genotype data, we tuned 30 different combinations of 4 hyperparameters separately for each psychiatric diagnosis (see Methods). The hyperparameter tuning results revealed diagnosis-specific patterns in the selection of top-performing combinations. For ADHD, the best-performing models consistently avoided kernel widths ≤ 32 in the locally connected layers (which determine how many input features each neuron processes), residual block dropout, which controls the probability of dropping connections in residual blocks to prevent overfitting values between 0.25 and 0.5, and the fraction of SNPs set to ‘missing’ in one-hot encoding for data augmentation set to 0.6 (**Figure S1A)**. In contrast, models predicting ASD and MDD were less sensitive to specific hyperparameter choices, achieving optimal performance across a broad range of combinations (**Figure S1B-C)**. For BIP, the top-performing models never selected kernel widths ≥ 64, a channel expansion base of 3 (which determines the number of trainable kernels applied to each local SNP region), a residual block dropout rate of 0.25, or a data augmentation setting where ≤ 20% of SNPs were set to ‘missing’ (**Figure S1D**). Similarly, for SCZ, the top-performing configurations consistently excluded a kernel width of 128 (**Figure S1D**). When testing for the impact that each hyperparameter had independently on the prediction performances, we found that the kernel width was significantly associated to higher AUROCs only for BIP (p-value 0.019), while the percentage of SNPs set to ‘missing’ negatively associated to lower AUROCs for ADHD and MDD with p-values of 0.017 and 0.039, respectively (**Figure S2, Table S1**). After 5-fold Cross Validation (5-CV) training in the iPSYCH1 cohort, the best performing combinations for each diagnosis resulted in average 5-CV AUROCs of 0.58, 0.56, 0.54, 0.55, 0.55 for ADHD, ASD, BIP, MDD, and SCZ, respectively. These findings highlight that the GLN architecture had varying degrees of sensitivity to hyperparameter selection across disorders, with some diagnoses exhibiting stricter constraints for optimal performance. Such differences may potentially reflect differences in the underlying genetic architecture of each disorder.

### GLN outperformed bigstatsr and generalized better on the replication set for ADHD, ASD, and MDD

To explore the potential advantages of using a DL model for computing *internal*, individual-based PGSs for psychiatric diagnoses, we compared the performance of the DL model GLN from the EIR framework to the model bigstatsr, a state-of-the-art linear model tailored to high dimensional individual-level genotype data (Privé et al., 2018, 2019). After training each model for each diagnosis using the same 5-fold CV splits, we found that bigstatsr consistently achieved higher performance than GLN on the iPSYCH1 held-out test set across all five diagnoses. However, these differences were not statistically significant, with average AUROCs ranging from 0.54 [0.49–0.58] for BIP to 0.61 [0.60–0.63] for ADHD (**Figure 2A, Table S4**). (R2s on liability scale from 0.004 [-0.003-0.011] for BIP and 0.035 [0.024-0.046] for ADHD, **Figure S4A, Table S4**). Nevertheless, when we evaluated the models on the iPSYCH2 replication set, GLN significantly outperformed bigstatsr in predicting ADHD, ASD, and MDD, with average increases in AUROC of 0.036 [0.025-0.046], 0.024 [0.012-0.036], and 0.02 [0.01-0.031], respectively (q-values, i.e. FDR corrected p-values < 0.001, **Figure 2A**, **Table S4**). We found this same pattern when comparing the performance in terms of R2, with average increases of 0.015 [0.01-0.02] for ADHD, 0.005 [0.003-0.007] for ASD, and 0.005 [0.002-0.008] for MDD (q-values < 0.001**, Figure S4A**, **Table S4**). To further assess the generalization performances of the models, we tested whether their performances on iPSYCH2 was lower than on the iPSYCH1 test set for each diagnosis. Here, bigstatsr exhibited a significant drop in AUROC performance for ADHD and MDD, with average decrease of 0.05 [0.035-0.07] and 0.04 [0.02-0.06], respectively (q-values < 0.001, **Figure 2B, Table S5**). In contrast, GLN showed no significant drop in performance for any diagnosis. In other words, it showed no signs of over-fitting to the training data. As before, we found the same pattern in terms of R2, with average drops of 0.027 [0.015-0.038] for ADHD and 0.015 [0.006-0.024] for MDD (q-values < 0.001, **Figure S4B, Table S5**). These results suggest that GLN produced more generalizable effects and the comparable performance of the two models within sample may be due to overfitting by bigstatsr.

**Figure 2.**
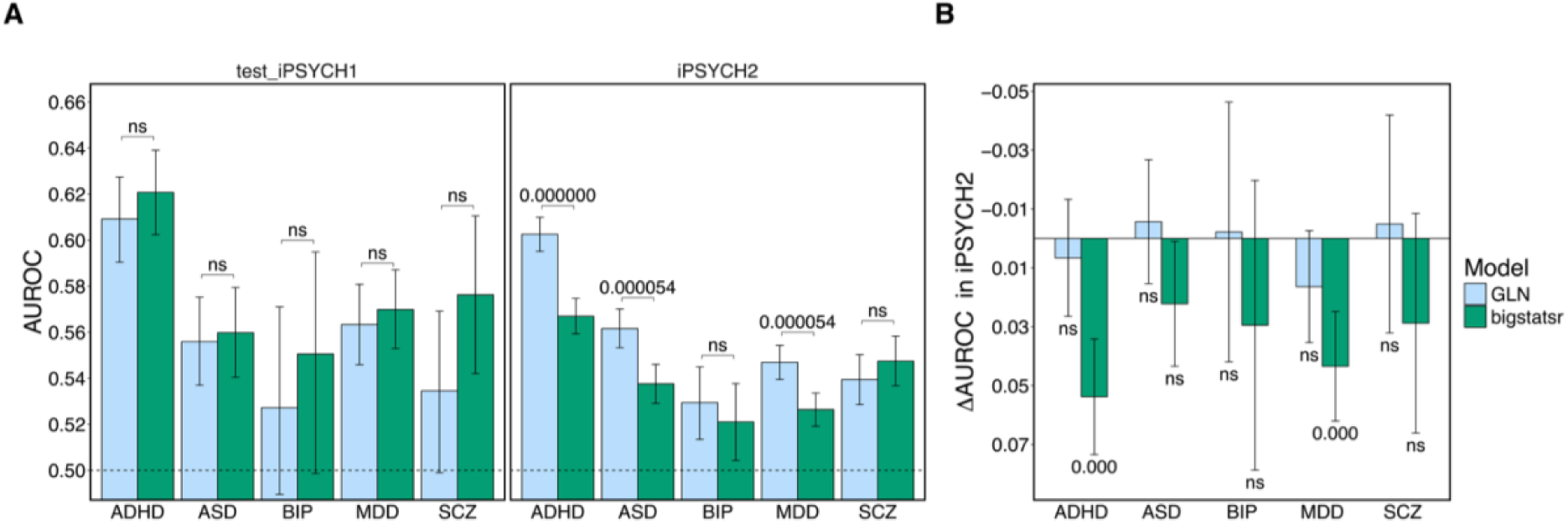
Performance and generalization of *internal* PGS models GLN and bigstatsr. **A)** Predictive performance in terms of AUROC of the two *internal* PGS models — GLN (DL-based, blue) and bigstatsr (linear, green) — for five psychiatric disorders (ADHD, ASD, BIP, MDD, SCZ) evaluated on the iPSYCH1 test set (left) and the out-of-sample iPSYCH2 replication set (right). Bars represent AUROC scores with error bars indicating 95% empirical confidence intervals based on 10,000 bootstraps. While the models showed comparable performance in-sample, GLN significantly outperformed bigstatsr for ADHD, ASD, and MDD on iPSYCH2 (FDR-corrected p-values annotated). **B)** Difference in AUROC (ΔAUROC) between iPSYCH1 and iPSYCH2 for each model, capturing drop in generalization performance. GLN exhibited more stable performance across cohorts, while bigstatsr showed significant performance declines for ADHD and MDD. Statistical significance was assessed using one-sided empirical tests with FDR correction; “ns” denotes non-significant comparisons.

### Integrating GLN and bigstatsr did not increase performance

Next, we explored whether a logistic regression-based integration of the prediction scores from GLN and bigstatsr could increase the classification performance compared to the models taken singularly. For each psychiatric disorder, we implemented a logistic regression model using the GLN and bigstatsr scores as explanatory variables. We trained each model on the same 5-CV folds used to train GLN and bigstatsr and evaluated the performance on the test set of the iPSYCH1 (see Methods). Here, the performance of the logistic regression integration models GLN+bigstatsr reached values comparable to those of GLN and bigstatsr alone, with AUROCs ranging from 0.54 [0.49-0.58] for BIP to 0.62 [0.60-0.64] for ADHD (**Figure 3A**, **Table S6**) (R2s from 0.003 [0-0.011] for BIP and 0.039 [0.028-0.051] for ADHD, **Figure S5A, Table S6**). Thus, the integrated model GLN+bigstatsr failed to provide a significant advantage neither in terms of AUROC, nor R2. This suggests that GLN and bigstatsr likely captured overlapping information and that their combination did not provide additional complementary signals to improve classification performance.

**Figure 3.**
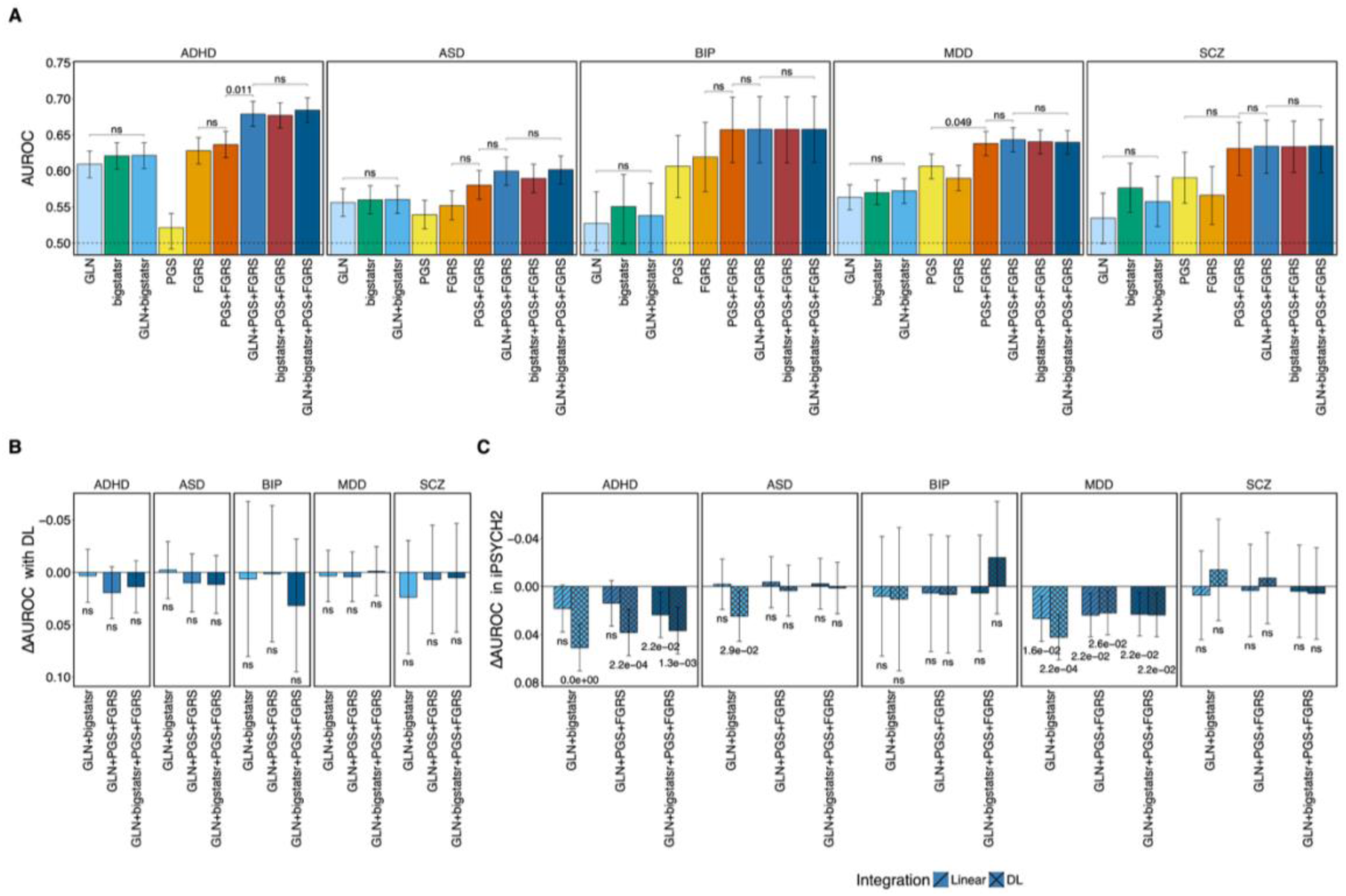
Integration of internal, external, and family-based scores improves ADHD prediction but shows limited benefit from DL-based combination. **A)** Predictive performance (AUROC) of different combinations of internal PGSs (GLN, bigstatsr), external PGSs, and family genetic risk scores (FGRSs) across five psychiatric disorders. For ADHD, the multivariate regression-based integration of GLN or bigstatsr with PGS and FGRS significantly improved prediction (GLN+PGS+FGRS AUROC = 0.68, *q* = 0.011), while other combinations showed no significant gains. For other disorders, individual and combined models performed comparably. **B)** Performance difference (ΔAUROC) between DL- and regression-based integration models across disorders. The DL-based integration did not outperform the regression-based one, with overlapping confidence intervals and no significant differences detected. **C)** Generalization performance (ΔAUROC between iPSYCH1 and iPSYCH2) for each integrated model. Regression-based integration methods (line pattern) were more robust to cohort differences, particularly for ADHD and ASD, while DL-based integration models showed larger generalization drops. Error bars represent 95% confidence intervals based on 10,000 bootstraps. Statistical significance was assessed using one-sided empirical tests with FDR correction; “ns” denotes non-significant comparisons.

### The *external* PGSs and PA-FGRSs exhibited similar performances for most disorders

We further investigated whether combining the *internal* PGSs – GLN and bigstatsr – with *external* PGSs and FGRSs could improve prediction performance. Since *external* PGSs leverage large-scale GWAS summary statistics and FGRSs capture family-based genetic risk, integrating them with logistic and/or DL-based *internal* PGSs could provide complementary predictive power. We first found that, when considered individually, PGSs and FGRSs exhibited similar predictive performance for ASD, BIP, MDD, and SCZ, with average AUROC values ranging from 0.55 [0.53–0.57] for ASD to 0.61 [0.56–0.66] for BIP (**Figure 3A**). The PGS for ADHD performed notably worse than both GLN and bigstatsr, as well as FGRS, with AUROCs of 0.52 [0.49-0.54] against 0.61 [0.60-0.63] and 0.63 [0.61-0.65], respectively (**Figure 3A**). This likely reflects the limited power of the external ADHD summary statistics since, unlike the other disorders in this study, most of the available genotype data of ADHD cases originate from iPSYCH, which was excluded from the training GWAS to avoid overfitting. The logistic regression combination of *external* PGSs and FGRSs (PGS+FGRS model) significantly outperformed the models with the individual scores for MDD but not for other disorders, reaching AUROC values of 0.64 [0.62-0.65] (empirical q-value 0.042, see Methods, **Figure 3A**). This was consistent with (Dybdahl Krebs et al., 2024), who observed that integrating PGS and FGRS scores increased MDD classification performance.

### Combining internal PGSs with PA-FGRS and external PGSs in a regression model improved classification performance in ADHD

We found that the multivariate model, which integrated the *internal* PGSs (GLN and/or bigstatsr) with *external* PGSs and FGRSs (GLN/bigstatsr+PGS+FGRS model), had significantly higher performance in terms of AUROC compared to the PGS+FGRS model for ADHD. Specifically, the AUROC reached 0.68 [0.66-0.70] for the multivariate model, compared to 0.64 [0.62-0.65] for the PGS+FGRS model (q-value 0.011, **Figure 3A**). This suggests that the *internal* PGSs provided additional information that was complementary to the *external* PGSs and FGRSs. However, when comparing the two models in terms of R^2^, the difference did not reach statistical significance but the direction of effect was the same, with the multivariate model reaching values of 0.081 [0.066-0.097] compared to 0.064 [0.050-0.078] for the PGS+FGRS model (**Figure S4**, **Table S7**). The *internal* PGSs GLN and bigstatsr likely enhanced classification by capturing information that improved case/control separation. However, these signals may explain only small incremental variance in ADHD liability, leading to a smaller change in R². This suggests that while *internal* PGSs improved case prediction in our setting, their contribution when combined with *external* PGSs trained on larger samples is limited. Finally, we found that a full multivariate model combining GLN+bigstatr+PGS+FGRS did not outperform the models that included only one *internal* PGS at a time, further suggesting that the two internal PGS methods GLN and bigstatsr provided similar weights to the same SNPs and thus did not provide complementary predictive value when combined (**Figure 3A**, **Table S7**).

### A DL-based integration of internal PGSs with PA-FGRS and external PGSs did not provide advantage over a logistic regression-based one

We further explored whether a DL based approach to integrating various combinations of risk scores would improve predictive performance compared to the regression-based integration methods tested above. In this DL-based strategy the GLN neural network underwent further training and combined the other scores in a way that allowed the detection of potential non-linear interactions between these and individual SNPs in the neural network architecture (see Methods). The DL-based integration models reached performances comparable to those of the regression-based integration models, with average AUROC differences between the DL and the logistic models ranging from -0.0025 [-0.0302 – 0.0251] for GLN+bigstatsr in ASD to 0.0321 [-0.0319 – 0.0961] for GLN+bigstatsr+PGS+FGRS in BIP (**Figure 3B**, **Table S8**). We observed the same pattern in terms of R2 (**Figure S5B**, **Table S8**). Thus, this analysis revealed no significant advantage of integrating *internal*, *external* PGSs and PA-FGRS using the DL approach over the regression-based one for any of the model combinations. These findings suggest that in this setting both the DL and regression-based integrations might capture overlapping genetic contributions to psychiatric disorders, implying a limited or absent contribution from complex and non-linear interactions among the tested set of predictors, or that in our scenario the DL models were unable to capture them.

### Generalization of all the models on iPSYCH2

Finally, we evaluated the replication performances of the logistic regression- and DL-based integration models on the iPSYCH2 out-of-sample set. We found that when predicting BIP and SCZ, none of the models had a significant drop in performance on iPSYCH2 with average differences in AUROCs between test iPSYCH1 and iPSYCH2 performances ranging from - 0.0239 [-0.0711 – 0.0233] for the DL model GLN+bigstatsr+PGS+FGRS in BIP to 0.0074 [- 0.0301 – 0.0448] for the logistic model GLN+bigstatsr in SCZ (**Figure 3C, Table S9**). This reflects the pattern we had already observed for the replication performance of the single *internal* PGS models GLN and bigstatsr (**Figure 2B**). However, the confidence intervals for BIP and SCZ were the widest among disorders studied here, since their sample sizes were the smallest (**Figure 1C)**. For MDD, all models had significant drops in performance, with average AUROCs differences ranging from 0.022 [0.004 - 0.04] for the DL model GLN+PGS+FGRS to 0.0425 [0.0237 - 0.0613] for the DL model GLN+bigstatsr (empirical q-values ≤ 0.015, **Figure 3C**, **Table S9**). This is consistent with the results observed for the single internal PGSs, where MDD showed the largest performance drop for GLN (average AUROC decrease of 0.0165 [–0.0022 – 0.0353] (**Figure 2B, Table S5**). As the GLN-based PGS for MDD had a poorer generalization performance compared to the other disorders, models that included it as part of the integration were similarly affected, leading to reduced overall performance. However, for ADHD and ASD, we found a clear pattern where the regression-based integration had a consistently lower drop in performance compared to the DL-based integration and was not significant for all models except GLN+bigstatsr+PGS+FGRS for ADHD (q-value 0.024, **Figure 3C**, **Table S9**). This suggests that the regression-based integrations were robust and able to better balance the good replication performances of GLN and the worse performances of bigstatsr.

## DISCUSSION

While efforts to refine polygenic scores have yielded substantial progress, predicting psychiatric disorders from genetics remains a challenge due to the complex genetic architecture of these conditions. Moreover, the extent to which DL models can offer improvements over simpler linear ones seems to currently be limited. Here, using the genotype data from the iPSYCH1 and 2 cohorts, we systematically evaluated two *internal* PGS methods for predicting five psychiatric disorders (ADHD, ASD, BIP, MDD, and SCZ): bigstatsr, a state-of-the-art linear model, and Genome-Local-Net (GLN), a deep learning (DL) model from the EIR framework (Privé et al., 2018, 2020; Sigurdsson et al., 2023). Additionally, we investigated whether integrating these *internal* PGSs with Pearson-Aitken Family Genetic Risk Scores (PA-FGRSs) and *external* PGSs, which use summary statistics for the estimation of the effect sizes, could improve predictive performance.

We found that the DL model GLN had a comparable performance to the linear model bigstatsr for predicting the five diagnoses on the held-out test set of the iPSYCH1 cohort. This is in agreement with previous work showing that DL and linear models can achieve similar performances for complex trait prediction from genetic data (Christodoulou et al., 2019; Ohta et al., 2024; Sigurdsson et al., 2023), especially if the genetic architecture of these traits is highly polygenic and thought to be mostly additive (Hill et al., 2005; Hivert et al., 2021). Bigstatsr is a highly tuned elastic net model which makes strong prior assumptions about the linear and additive effects of the SNPs, and this in turn makes it perform well for traits thought of as being mainly additive (Momen et al., 2018). Conversely, DL models like GLN inherently make fewer assumptions about the distribution and nature of the effect of genetic variants compared to traditional linear models. This reduced assumption framework complicates the training process and necessitates larger sample sizes for effective model training (Grealey et al., 2024).

Interestingly, GLN had a higher generalization performance compared to bigstatsr when applied to the out-of-sample replication set of iPSYCH2 for predicting ADHD, ASD, and MDD, both in terms of AUROC and R2. This finding suggests that the DL model GLN was able to capture more robust, potentially non-linear and/or interaction-based genetic patterns underlying the psychiatric disorders, while the linear model bigstatsr might have been more prone to overfitting to the specific characteristics of the initial training cohort. We previously showed that the GLN model exhibited robust generalization performance across two independent cohorts (Sigurdsson et al., 2023). However, that study did not focus on psychiatric disorders nor included a comparison with bigstatsr, preventing us to directly compare our results. While GLN’s superior performance in the replication set suggests better generalization, we cannot exclude that this advantage arose from the model’s ability to capture subtle patterns specific to the training data, which coincidentally aligned with the replication set, instead of capturing real and more generalizable genetic effects. Further research on this and potentially other cohorts would be necessary to disentangle this effect and validate GLN’s increased generalizability for psychiatric disorder prediction. If this finding holds, it will undoubtedly represent an interesting avenue of research since classic PGSs generally suffer from poor transferability across cohorts (Ding et al., 2023; Wang et al., 2022).

Combining the scores from GLN and bigstatsr did not provide additional predictive performance compared to the single models, suggesting that these two *internal* PGS methods might have captured largely redundant genetic effects in the training set of the iPSYCH1 cohort. In order to confirm this hypothesis, future analyses could examine the specific genetic variants to which the two models assigned their weights and assess the extent of their overlap.

We further showed that integrating either GLN or bigstatsr with *external* PGS and FGRS in a logistic model significantly increased the prediction performance for ADHD but not for the other disorders. This could be an indication that the ADHD *internal* PGSs - GLN or bigstatsr - were able to capture information from the genetic data beyond what was obtained through the external PGS and FGRS. While this improvement was significant in terms of AUROC, it did not reach significance when evaluated using R² on the liability scale. However, the trend in R² remained in the same positive direction, indicating a consistent, albeit more modest, gain. AUROC reflects a model’s ability to rank cases above controls and is thus sensitive to improvements in classification, whereas R² measures the proportion of variance explained in the underlying liability. Since PGSs tend to perform best in identifying individuals at the extremes of the risk distribution (Choi et al., 2020), it is possible that the internal PGSs improved classification by better identifying these individuals, which enhanced AUROC but had a smaller impact on overall explained variance.

Previous studies have showed how combining multiple risk scores can increase the performance for predicting psychiatric disorders and other complex traits (Albiñana et al., 2023; Allesøe et al., 2022; Dybdahl Krebs et al., 2024; Elgart et al., 2022; Truong et al., 2024). However, to the best of our knowledge, this is the first study that explored the potential of combining scores using DL and combining both *internal* and *external* PGSs, together with FGRSs, to predict psychiatric disorders. Even though the performance of the full logistic model combining all these risk scores was significantly higher than the one of the PGS+FGRS model for ADHD, the increase was modest. In addition, the DL-based integration did not provide significant advantages over the logistic one. The role of non-linear interactions in complex traits remains a subject of ongoing research. Here we did not observe substantial performance differences between deep learning models and linear approaches for psychiatric disorders. This could reflect a genuine lack of non-linear signal, or it may be due to limitations in our current sample size and experimental setup, which may not have been sufficient to detect such effects (Grealey et al., 2024). Nonetheless, we observed indications of improved generalization of the DL models, though these findings require further validation. The difficulty in detecting evidence of non-linear interactions in complex human traits may not be due to their absence but rather due to the multitude and subtlety of the effects. Consequently, linear approximation could manage to capture nearly all the population-level variation.

## LIMITATIONS

Our study has several limitations. A first and inherent limitation is the sample size which, while being the largest for traits like ADHD and ASD, may still be insufficient for the DL model to fully capture the complex genetic effects of these disorders. As sample sizes increase, the relative performance difference between DL and linear models is envisaged to change (Grealey et al., 2024). Another limitation that may have impacted the performance of the DL model is the computational constraint, which restricted us to testing only a limited number of hyperparameter combinations. A more exhaustive hyperparameter optimization or model search could potentially enhance the performance of the DL model. However, the DL model’s optimization process is more challenging due to the higher number of potential hyperparameters compared to the bigstatsr model. Additionally, for computational feasibility and fair comparisons we restricted the genotype data to a relatively small number of SNPs, which may mask some potential of the DL models. In contrast, bigstatsr can handle a larger number of SNPs more efficiently, allowing for faster processing and analysis (Privé et al., 2018, 2020). Related to this, another limitation is the specific architecture of the GLN model, which is not representative of all DL models. Therefore, we cannot assume these results would generalize to all other architectures. In contrast, linear models have fewer variations in their design and implementation, making them more straightforward to compare. As a result, our findings regarding the performance of the GLN model should not be generalized to all DL approaches. Finally, since we focused on a cohort with a homogeneous European ancestry, we did not consider the remaining potential biases due to population structure, and we cannot exclude with certainty that it would not impact the differences between the models presented in this study.

## CONCLUSIONS AND FUTURE PERSPECTIVES

In this study we compared a DL model, Genome-Local-Net (GLN), with the linear model bigstatsr for predicting five psychiatric disorders using genotype data from the iPSYCH cohort. While both models showed similar performance within the training cohort, GLN generalized better to an independent replication set for ADHD, ASD, and MDD. This suggests that deep learning may offer advantages in terms of generalizability, although its benefit was not consistent across all disorders. Integrating these *internal* polygenic scores with *external* GWAS-based PGSs and FGRS improved prediction for ADHD, but not for the other disorders. Notably, logistic regression-based integration performed as well as or better than deep learning-based integration, indicating that complex interactions among scores were either absent or not effectively captured by the deep models.

In conclusion, our findings suggest that DL approaches can be a valuable tool for complex trait prediction from genetic data, with the potential for improved generalizability compared to linear models. However, the performance gains may be limited, at least at current sample sizes, for highly polygenic traits with a mostly additive genetic architecture, such as the psychiatric disorders studied here. Further research is necessary to explore the advantages of deep learning for other complex traits, as well as to investigate more effective ways of integrating various genetic risk factors using machine learning. Overall, the clinical utility of these models is currently limited, but they hold great promise for future clinical deployment as sample sizes increase and our understanding of the genetic underpinnings of these disorders continue to advance. Multi-modal models, which integrate diverse data sources such as genetic, environmental, and clinical information, are already being used in research to provide a more comprehensive study of complex traits (Allesøe et al., 2022; Ma et al., 2025; Porcu et al., 2021; Sigurdsson et al., 2024). In the future, we anticipate an increasing adoption of these models, as they will not only enhance the study of genetic factors but also incorporate environmental influences, allowing for a more holistic perspective on their interactions and overall impact.

## MATERIALS AND METHODS

### Dataset

The dataset used in this study originates from the Lundbeck Foundation Integrative Psychiatric Research (iPSYCH) consortium, which created the iPSYCH2015, a large case-cohort sample encompassing the entire population of Denmark born between 1981 and 2008 (Bybjerg-Grauholm et al., 2020; Pedersen et al., 2018). From this population of 1,657,499 citizens, 88,831 individuals were selected and genotyped based on being diagnosed with at least one major mental disorder, and 48,420 individuals were randomly selected as controls from the general population, for a total of 137,251 individuals. The iPSYCH2015 case-cohort sample is comprised of two separate enrollment periods based on the initial population - iPSYCH2012 and iPSYCH2015i, hereafter referred to as iPSYCH1 and iPSYCH2 for simplicity. The first includes 57,377 psychiatric cases and 30,000 random population controls, totaling 86,189 individuals. The second enrollment period expanded the sample size by adding 36,741 psychiatric cases and 19,982 random population controls, for a total of 56,233 individuals. All the individuals in the cohort got a blood sample taken at birth and stored in the Danish Neonatal Screening Biobank (Nørgaard-Pedersen & Hougaard, 2007). From these, DNA was extracted, and genotyping was performed on the Infinium PsychChip v1.0 array for iPSYCH1 and on the Global Screening Array v2 for iPSYCH2. The steps of genotyping, quality controls and imputation have been described in detail in Schork et al., 2019. Only unrelated individuals with European ancestry were included in this study. To ascertain psychiatric diagnoses, the iPSYCH study design relied on data retrieved from the Danish Psychiatric Central Research Register (PCR)(Mors et al., 2011), in conjunction with information sourced from the Danish National Patient Register (DNPR) (Lynge et al., 2011). The diagnoses were administered by psychiatrists during in and out-patients encounters up until 2016. Diagnoses from primary care were not registered in the national registry and therefore not available for this study. The mental disorders included in the iPSYCH cohort and which we analyzed in this study were defined according to the International Statistical Classification of Diseases and Related Health Problems, Tenth Revision (ICD-10) and are the following: attention-deficit/hyperactivity disorder (ADHD: F90.0), autism spectrum disorders (ASD: F84.0, F84.1, F84.5, F84.8, F84.9), bipolar disorder (BIP: F30-31), major depressive disorder (MDD: F32, F33), and schizophrenia (SCZ: F20). The personal information of the individuals enrolled in the study was anonymized to ensure their privacy. The project received approval from the Danish Data Protection Agency and, therefore, under Danish law, obtaining informed consent from the participants was not necessary.

### Genotype data preprocessing

To ensure a computationally feasible size of the data, we selected SNPs in an informed manner, leveraging GWAS summary statistics for each disorder to define the final list of input variants. An overview of the filtering steps is presented in **Figure 1C**. We filtered the genotype data from the iPSYCH1 and iPSYCH2 cohorts independently to remove multi-allelic variants, and to retain only variants with a minor allele frequency (MAF) > 0.01 and with an imputation quality score (DR2) > 0.8. The preprocessing steps were carried out with custom scripts using bcftools (v.1.20) and PLINK (v.2.00) (Chang et al., 2015; Danecek et al., 2021). We then took the intersection of the variants from the two cohorts, resulting in 6,844,196 SNPs. To construct a GWAS-informed SNP set, we ran clumping and thresholding (C+T) with PRSice (v.2.3.5) (Choi & O’Reilly, 2019) on each of the 5 psychiatric disorders ADHD, ASD, BIP, MDD, SCZ, and across a range of p-value thresholds: 0.00000005, 0.0000001, 0.000001, 0.00001, 0.0001, 0.001, 0.01, 0.05, 0.1, 0.2, 0.3, 0.4, 0.5, 0.6, 0.7, 0.8, 0.9, 1 with the options --`clump-kb 250 –clump-p 1 –clump-r2 1`. This was performed on the training and validation set of iPSYCH1 and not on the entire cohort to prevent data leakage. This resulted in 18 different SNP sets for each psychiatric disorder, one for each p-value threshold from the C+T procedure. For each of these SNP sets at each p-value threshold, we then took the union of the SNPs across the 5 disorders and further clumped these candidate SNP sets using the function `snp_clumping` from the package bigsnpr (v.1.11.6) (Privé et al., 2018) across a range of R2: 0.1, 0.2, 0.3, 0.4, 0.5, 0.6, 0.7, 0.8, 0.9 and using MAF as a statistic, to ensure that no genomic region was left unrepresented (Privé et al., 2018). From these, we selected the final SNP set as the one having 230,588 SNPs, obtained using a p-valued threshold of 0.05 and a clumping R2 of 0.5. This was an arbitrary choice, based on a tradeoff between selecting a feasible number of variants with enough information but at the same time with reduced LD. Only the genotype data from the autosomal chromosomes was used in this study. The overview of the number of variants in each SNP set created to select the final set is shown in **Table S10**.

### Implementation of training

For each mental disorder, we split the iPSYCH1 dataset into training and test sets with ratios of 0.9 and 0.1, respectively, stratifying for case/control status. The test sets were held out and used exclusively for the evaluation of the models. The training sets were further split into 5 folds so that we could train the models in a 5-fold cross-validation fashion. The same folds were used for all models to ensure reproducibility and consistency in their comparisons. The final split sizes (train/validation per fold/test) were the following: ADHD: 34,681/6,936/3,854, ASD: 32,692/6,539/3,633, BIP: 23,412/3,682/2,602, MDD: 38,106/7,622/4,234, and SCZ :24,329/4,886/2,704. The final models were the result of an ensemble of the five models trained on the five different training folds and evaluated on the test set and on the entirety of the iPSYCH2 dataset as an out-of-sample replication set.

### GLN implementation and hyperparameter tuning

The neural network models used in this study employ the genome-local-net (GLN) architecture developed in (Sigurdsson et al., 2023). In this architecture, locally connected layers (LCL) and residual blocks were employed to extract information from the genotype data. This allowed for the combination of both local and global genomic information while ensuring that the number of parameters was kept manageable compared to using other architectures such as fully connected (FC) layers. The models were implemented using EIR (v.0.1.39). The genotype data was one-hot encoded allowing each allele to have a distinct weight. For each of the five psychiatric diagnoses, we performed hyperparameter tuning with a random search approach on the training and validation set of iPSYCH1. We considered four hyperparameters for tuning. The kernel width, which determines the receptive field size in locally connected layers, was tested at values of 16, 32, 64, and 128 (covering 4, 8, 16, and 32 one-hot encoded SNPs, respectively). The residual block dropout, which controls the probability of dropping connections in residual blocks to prevent overfitting, was evaluated at dropout rates of 0, 0,10, 0.25, 0.5, and 0.9. The channel expansion base, which defines the power of 2 used to determine the number of channels (i.e., number of trainable kernels applied to each SNPs region) in the network, was tested with values of 2, 3, and 4. Finally, the percentage of SNPs set to ‘missing’ for data augmentation, a method that simulates missingness in genotype data by randomly setting a proportion of SNPs to ‘missing’ in one-hot encoding, was explored at levels of 0%, 20%, 40%, and 60%. Each hyperparameter tuning training was performed in a 1-fold fashion, using validation sets 2-5 for training and validation 1 for evaluation and selection of the best hyperparameter combinations based on the AUROC metric.

### Linear model bigstatsr

The bigstatsr model was trained using the same 5-CV folds as used for the training of the GLN model with the function `big_spLogReg` from the bigstatsr and bigsnpr packages (v1.5.12 and v.11.6, respectively) (Privé et al., 2018, 2020). This function natively implements a Cross-Model Selection and Averaging (CMSA) method, which enables a fast cross-validation coupled with an extensive and automatic hyperparameter tuning of the lambda and alpha hyperparameters of the elastic net regularization. We performed a grid search for alpha with values [0.0001, 0.001, 0.01, 0.1, 1] and we used the default options to select the best penalization parameter (λ). The final model was a result of the average of the 5 models run on each fold, as an ensemble model.

### *External* PGSs computation

We computed *external* polygenic scores (PGSs) for the individuals of the two cohorts iPSYCH1 and iPSYCH2 separately, using summary statistics from recent genome-wide association studies (GWAS) conducted by the Psychiatric Genomics Consortium (PGC) on ADHD (Demontis et al., 2023), ASD (Grove et al., 2019), BIP (Mullins et al., 2021), MDD (Howard et al., 2019), SCZ (Trubetskoy et al., 2022).To avoid sample overlap, we ensured that iPSYCH participants were excluded from the discovery datasets. We re-estimated SNP effect sizes using SBayesR from GCTB (v.2.05beta) (Lloyd-Jones et al., 2019), a Bayesian multiple regression method, applying a p-value threshold of 0.9. For linkage disequilibrium (LD) modeling, we used the HapMap3-based banded LD matrix, downloaded from the GCTB resource (https://cnsgenomics.com/software/gctb/#Download). We used the default SBayesR settings for the scaling factor (γ = 0, 0.01, 0.1, 1) and mixture proportions (π = 0.95, 0.02, 0.02, 0.01). After rescaling the SNP effects, we generated PGS for each individual using the scoring module in PLINK (v.2.00) (Chang et al., 2015).

### PA-FGRSs

The PA-FGRS were computed as previously described (Dybdahl Krebs et al., 2024) using the PA-FGRS R package (https://github.com/BioPsyk/PAFGRS). In summary, we used the Danish Civil registry to link each member of the iPSYCH study to all available relatives (21 on average). For each relative diagnoses were extracted from PCR and DNPR and date of end-of-follow-up was set the date of death, emigration or Dec 31, 2016, whichever came first. Assuming an additive genetic liability threshold model, the PA-FGRS method estimates the expected genetic liability of an individual conditional on the disease status of all available relatives, while modeling their relatedness, the sex-stratified disease prevalence and proportion their life they have been observed which is integrated using the sex- and birthyear specific cumulative incidence curves.

### Logistic regression-based integration

For each mental disorder, we carried out an integration of the scores from GLN, bigstatsr, *external* PGS and FGRS using a logistic regression and implemented using the tidymodels framework in R. For the ADHD, ASD, and MDD models, we allowed an interaction term between the scores, while we did not for the BIP and SCZ models, since it negatively impacted the performance in terms of AUROC (**Fig S6**, upper plots). As mentioned above, the training was performed on the same 5-CV folds as for the other models, and the performance was evaluated on the test sets of iPSYCH1 and the out-of-sample replication sets of the iPSYCH2. To avoid data leakage, the scores were centered and scaled using the means and standard deviations calculated on the training set.

### DL-based integration

We performed the DL-based integration of the GLN model with the scores from bigstatsr, *external* PGS and FGRSs, by allowing the pre-trained GLN model to undergo further training and by adding the scores as tabular input to EIR. Initially, the GLN model was trained solely on the genotype data. The scores from bigstatsr, *external* PGSs and FGRSs were then fed as additional inputs to this model, via a dedicated tabular feature extractor. This used embeddings for the categorical inputs, and the continuous inputs where normalized with the means and standard deviations computed from the training set to avoid data leakage. The components of the pre-trained GLN model (genotype feature extractor, fusion module and output) were kept the same. The new tabular inputs were injected into the existing fusion module through the tabular extractor, enabling a flexible architecture that could potentially learn interactions between individual SNPs and tabular scores (e.g., a specific SNP modulating the effect of a PGS). A schematic overview of the architecture is available in Figure 2A from the original EIR paper (Sigurdsson et al., 2023). All models allowed for interaction between the scores, since for the DL case the performance of these models was not significantly lower than those of the non-interaction models for BIP and SCZ (**Fig S6**, bottom plots)

### Model performances and statistical tests

For each psychiatric disorder, we trained all models – GLN, bigstatsr, the regression- and the DL-based combined models of all scores – as five separate models using five-fold cross validation. We then formed the final models by ensembling these five trained models with a simple averaging of the predictions of each model. We used these ensemble models to predict the mental disorders on the iPSYCH1 tests sets, as well as on the full iPSYCH2 samples, which served as an out-of-sample replication set. To derive confidence intervals for the performance metrics and estimate their sampling distributions, we bootstrapped the test and replication sets, i.e. for each set we drew 10,000 samples with replacement with the same original size, and computed the average metric, together with its standard deviation and non-parametric 2.5^th^ and 97.5^th^ percentiles (Raschka, 2020). To compute the Area Under the Receiver Operating Curve (AUROC), we used the same implementation of the AUCBoot function from the bigstatsr package (Privé et al., 2020). For the R2, we used the same approach as (Albiñana et al., 2023), namely we computed the R2 using the caret function R2 on the predicted probabilities and true outcomes, and then converted this onto the liability scale using the function coef_to_liab from the bigsnpr package (Privé et al., 2018) using the following values as prevalences for the psychiatric disorders: ADHD: 0.05, ASD: 0.02, BIP: 0.01, MDD: 0.15, SCZ:0.01. For the option K_gwas of the function, analogously to what (Albiñana et al., 2023) did, we used the proportion of cases in the specific sets (these are shown in **Figure 1C**). We compared the performance of two models by deriving empirical p-values using the bootstrapped AUC and R2 for each model, aligning with the classical definition of a p-value as the probability, under the null hypothesis, of observing data at least as extreme as the observed differences. We performed a one-side empirical test to assess whether e.g. Model 2 achieved higher performance than Model 1, by counting the proportion of times the metric from Model 2 was higher or equal than the metric from Model 1. Under the null hypothesis of “Model 2’s metric is not higher than Model 1’s metric”, this proportion served as the p-value for the one-sided test. To correct for multiple comparisons, we applied the false discovery rate (FDR) method using the Benjamini–Hochberg procedure(Benjamini & Hochberg, 1995). This approach provides adjusted p-values, often referred to as “q-values,” which control the expected proportion of false positives (Type I errors) across all comparisons performed, and was applied with the function `p.adjust` from the stats package in R.

## Data Availability

iPSYCH data are stored at a secure national HPC facility in Denmark. The iPSYCH consortium supports data access for researchers under Danish legal frameworks. Although the data used in this study is not publicly available, due to restrictions on sensitive person-level data, access may be granted upon request to the iPSYCH consortium.

https://github.com/leocob/eirpsy

## DATA AND CODE AVAILABILITY

The iPSYCH data is securely stored and was analyzed in the national HPC facility in Denmark (https://genome.au.dk/). The personal data was anonymized prior to use for research purposes. The project received approval from the Danish Data Protection Agency, and in accordance with Danish legislation, informed consent was not required. The iPSYCH initiative is committed to providing access to these data to the scientific community, in accordance with Danish law. EIR is available at https://eir.readthedocs.io/en/stable/. The code used in this project is available at https://github.com/leocob/eirpsy. The code to compute PA-FGRSs is available at https://github.com/BioPsyk/PAFGRS.

## SUPPLEMENTARY INFORMATION

Supplementary figures and tables are available in the attached files.

## AUTHOR CONTRIBUTIONS

Conceptualization: S.R, A.J.S, M.E.B. Data curation: L.C. Formal Analysis: L.C. Investigation: L.C. Methodology: L.C., A.I.S Resources: M.V., K.L.G.H., M.D.K., M.E.B., iPSYCH Study Consortium Software: L.C., A.I.S Supervision: A.J.S, S.R., A.I.S, J.M., M.E.B. Visualization: L.C. Writing – original draft: L.C. Writing – review & editing: L.C., A.J.S, S.R., M.E.B., T.W.

## ACKNOWLEDGEMENTS

We would like to thank Professor Ditte Demontis for providing us with the latest GWAS summary statistics for ADHD trained on PGC excluding the iPSYCH individuals.

