## Supplementary Figures for "Deep learning-based polygenic scores enhance generalizability of psychiatric disorders prediction"

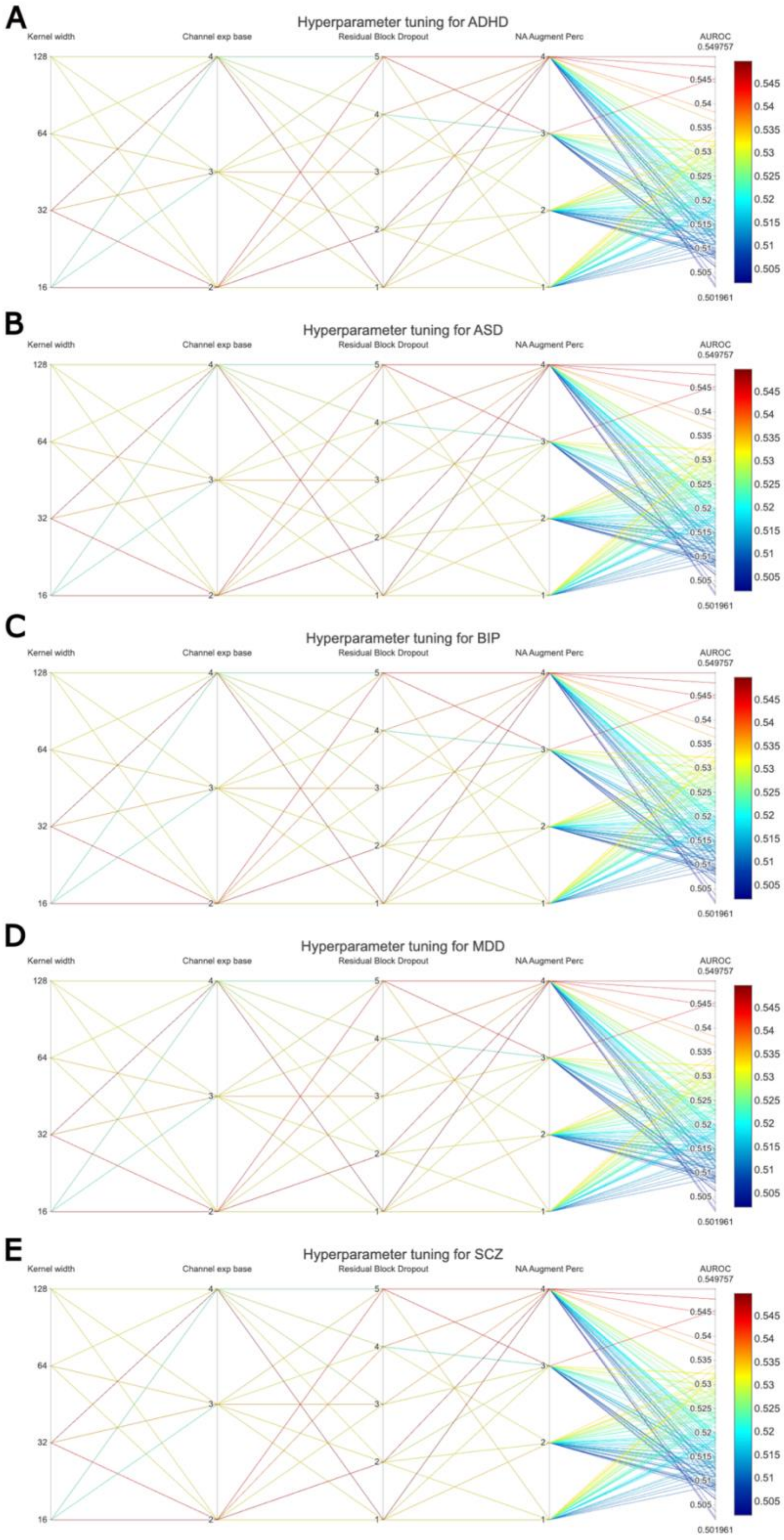


**Figure S1. Hyperparameter tuning results for the GLN deep learning model across psychiatric disorders.** Parallel coordinate plots showing the impact of different hyperparameter combinations on model performance (measured by AUROC) for each psychiatric disorder: ADHD, ASD, BIP, MDD, and SCZ. Each line represents a distinct combination of four hyperparameters: kernel width (receptive field size), channel expansion base (impacting network width), residual block dropout rate (*rb_do*), and percentage of SNPs set to missing for data augmentation (*na_augment_perc*). Line color indicates the resulting AUROC, with warmer colors representing higher performance. Diagnosis-specific trends emerged—for example, higher AUROCs for ADHD were associated with kernel widths > 32 and dropout rates between 0.25 and 0.5, while BIP showed consistently poorer performance with larger kernel sizes (≥ 64). These findings illustrate that GLN's optimal performance is sensitive to hyperparameter selection and varies across psychiatric traits.

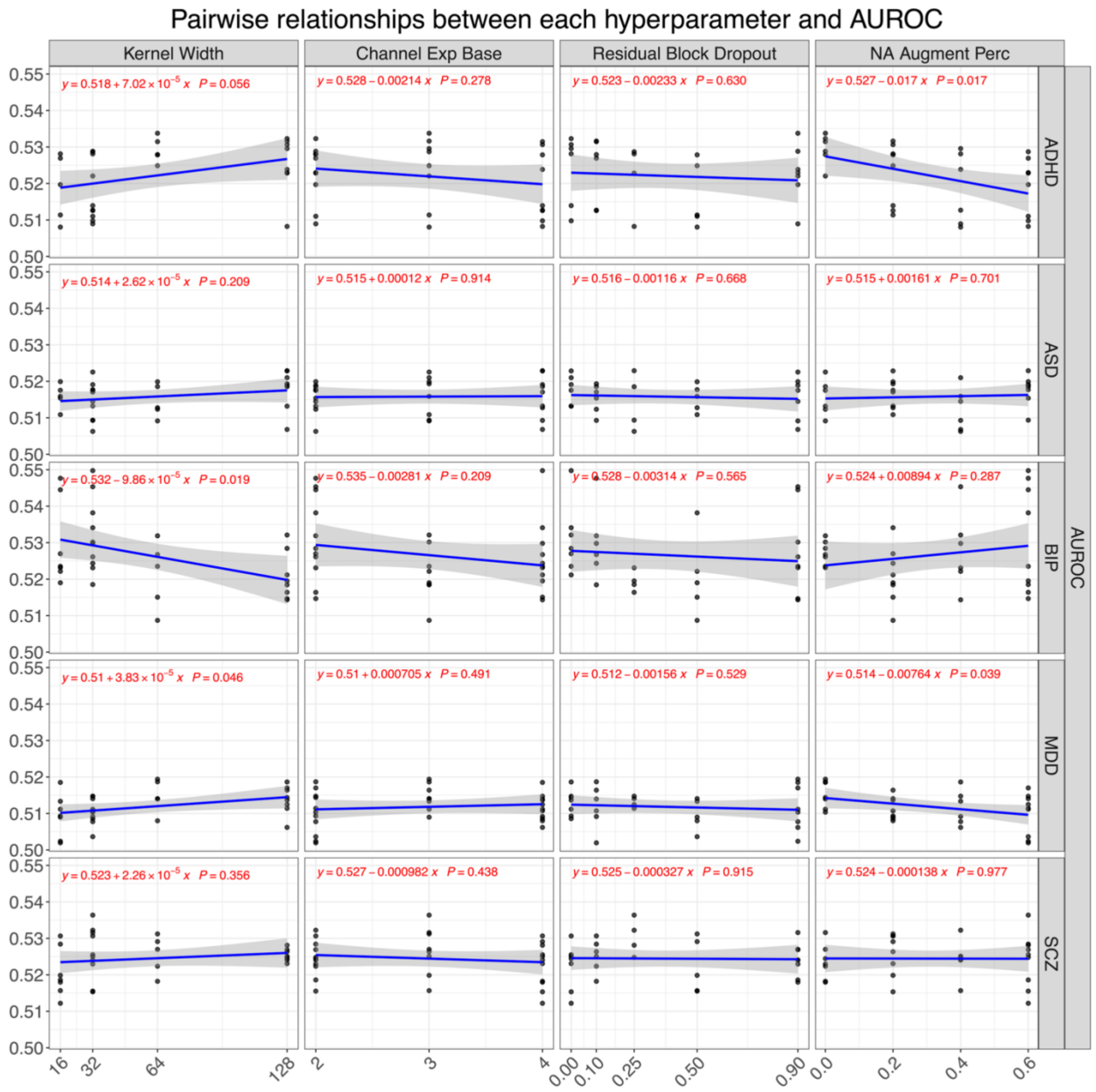


**Figure S2. Effect of individual GLN hyperparameters on predictive performance across psychiatric disorders.** Linear regression plots showing the pairwise relationship between each of the four hyperparameters used in the GLN model - kernel width, channel expansion base, residual block dropout (*rb_do*), and percentage of SNPs set to missing for data augmentation (*na_augment_perc*) - and AUROC for each psychiatric disorder. Each dot represents one hyperparameter combination, and the fitted blue line with gray confidence band represents the regression trend. Red equations and *P*-values denote the slope and statistical significance of each relationship. Notably, larger kernel widths were significantly associated with decreased performance for BIP ($P = 0.019$), while higher missingness rates were significantly associated with reduced AUROC for ADHD ($P$ = 0.017) and MDD ($P$ = 0.039). These results highlight that specific hyperparameters may exert diagnosis-specific effects on model performance.

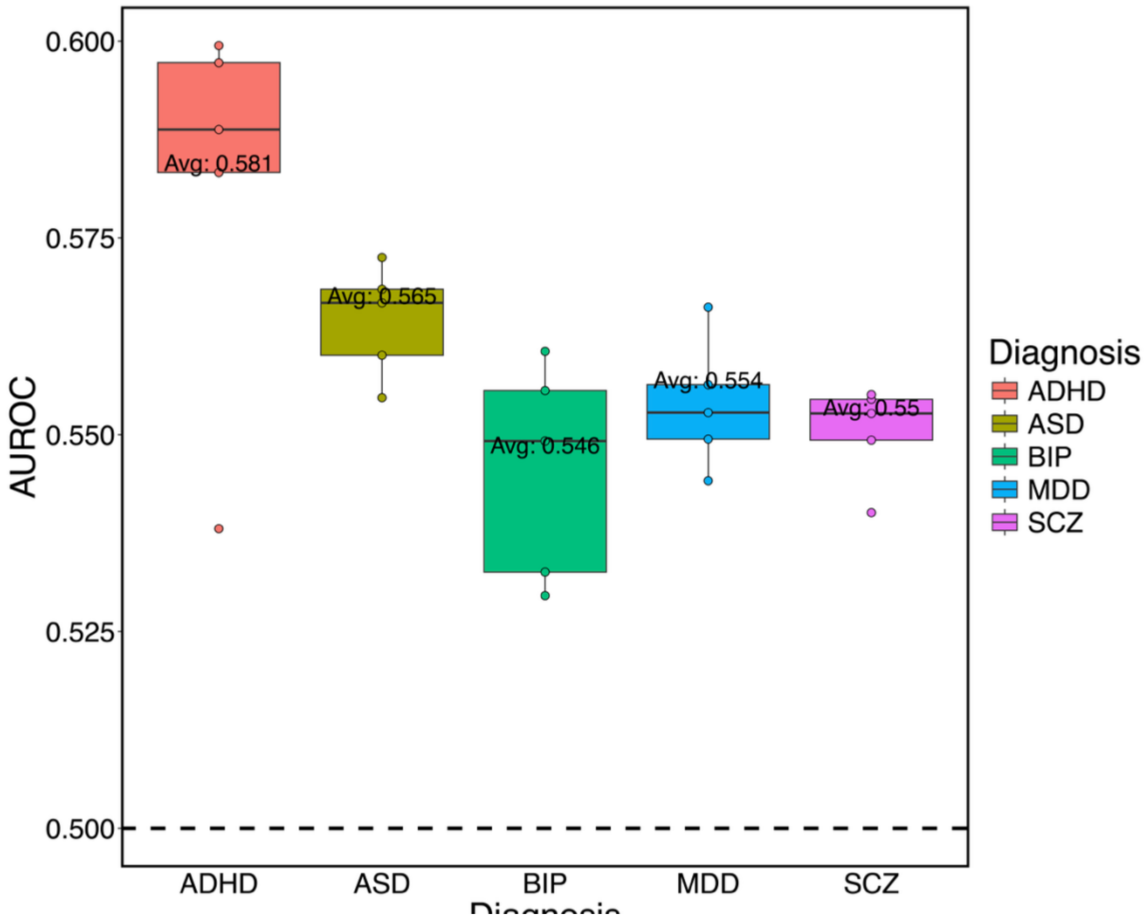


**Figure S3. Distribution of cross-validated AUROC scores for GLN across psychiatric disorders.** Boxplots showing the distribution of AUROC values obtained from 5-fold cross-validation of the best-performing GLN hyperparameter combinations for each psychiatric disorder in the iPSYCH1 training set. Each box represents the variability in AUROC across folds, with the average AUROC annotated. ADHD achieved the highest average predictive performance (AUROC = 0.581), followed by ASD (0.565), MDD (0.554), BIP (0.546), and SCZ (0.550). These results illustrate diagnosis-specific differences in model performance, with ADHD exhibiting more consistent and higher classification accuracy relative to the other disorders.

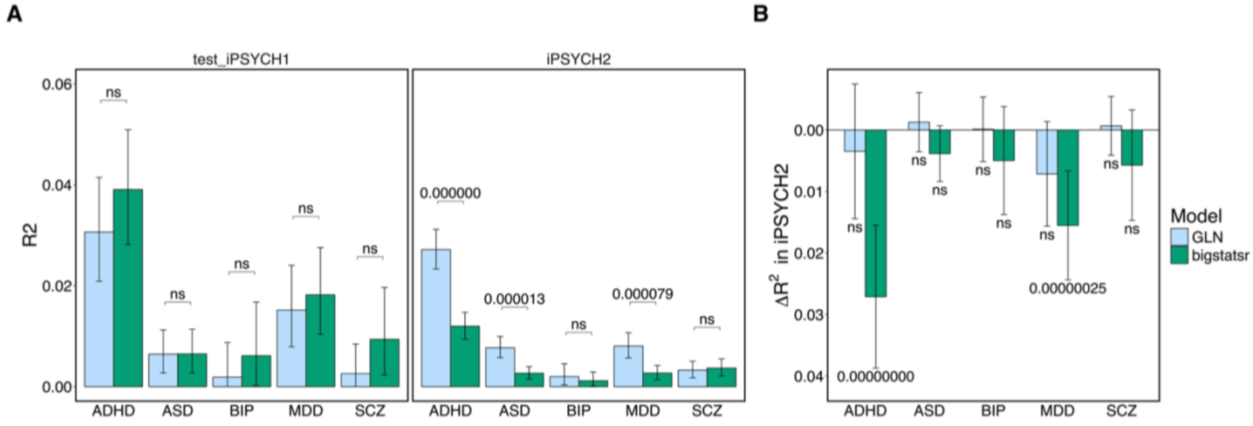


**Figure S4: Performance and generalization of internal PGS models GLN and bigstatsr in terms of R2. A)** Performance in terms of coefficient of determination ($R^2$ on the liability scale) for GLN (light blue) and bigstatsr (green) models across five psychiatric disorders in the iPSYCH1 test set (left) and iPSYCH2 replication set (right). While in-sample performance was similar across models, GLN showed significant higher $R^2$ than bigstatsr for ADHD, ASD, and MDD in iPSYCH2 (FDR-corrected q-values indicated). **B)** Generalization performance assessed via the change in $R^2$ ($\Delta R^2$) between iPSYCH1 and iPSYCH2. For ADHD and MDD, bigstatsr showed significant drops in $R^2$, while GLN maintained more stable performance. Error bars denote 95% confidence intervals based on 10,000 bootstrap estimates. These findings mirror AUROC-based results and support GLN's improved robustness in out-of-sample prediction for select disorders.

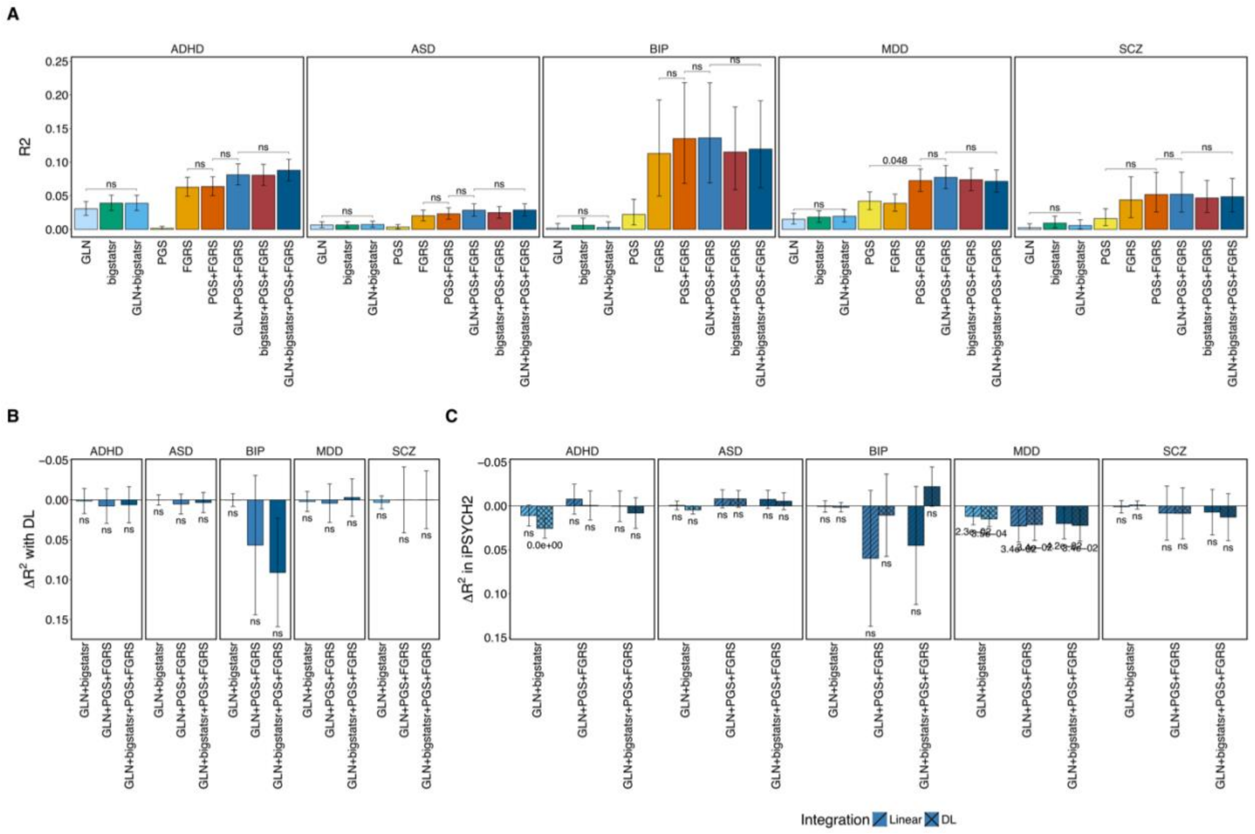


**Figure S5. R²-based evaluation of integration models across disorders and cohort. A)** R² (on the liability scale) of different model combinations for each psychiatric disorder in the iPSYCH1 test set. Models include *internal* PGSs (GLN, bigstatsr), external PGSs, family genetic risk scores (FGRSs), and their linear integrations. **B)** Difference in R² (ΔR²) between DL-based and linear integration models, showing that DL combinations did not significantly outperform their linear counterparts across any disorder. **C)** Generalization performance (ΔR² between iPSYCH1 and iPSYCH2) for each integration model. DL-based integrations showed significant performance drops for ADHD and ASD. Linear models were generally more stable across cohorts. Error bars represent 95% confidence intervals estimated via 10,000 bootstraps.

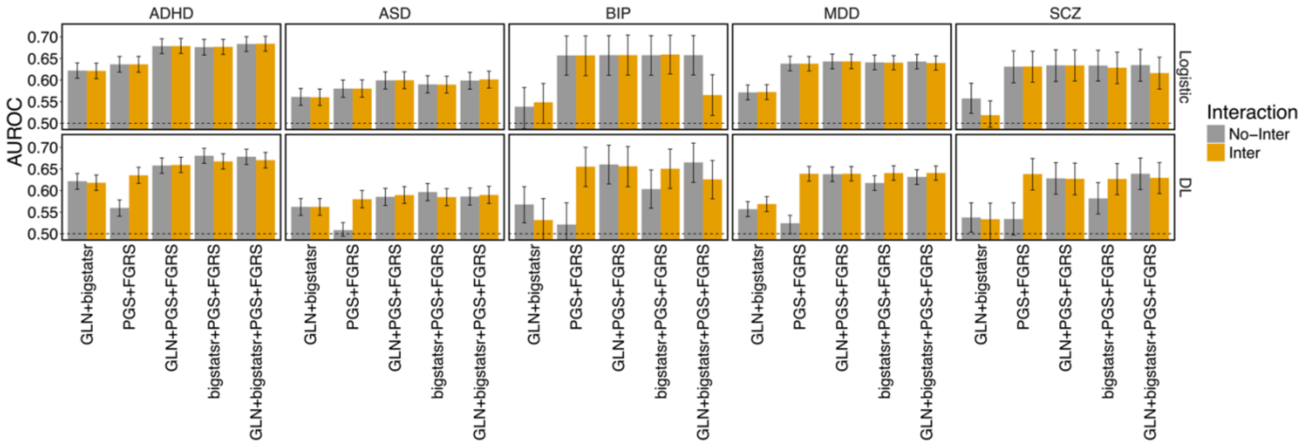


**Figure S6. Impact of interaction terms on model performance across disorders and integration strategies.** Bar plots showing the AUROC of integration models with and without interaction terms across ADHD, ASD, BIP, MDD, and SCZ. Each model includes different combinations of *internal* (GLN, bigstatsr), *external* (PGS), and family-based (FGRS) risk scores, integrated using either a linear approach (top) or a DL-based one (bottom). Models without interaction terms are labelled "No-Inter" (gray) and those with are labelled "Inter" (orange). For linear models, including interactions tended to reduce performance for BIP and SCZ, while DL-based models were less affected by the inclusion of interactions. These results guided the decision to exclude interaction terms in BIP and SCZ linear models in the main analysis.
